# Machine learning for identifying relevant publications in updates of systematic reviews of diagnostic test studies

**DOI:** 10.1101/2020.06.16.20132670

**Authors:** Toni Lange, Guido Schwarzer, Thomas Datzmann, Harald Binder

## Abstract

**Background:** Updating systematic reviews is often a time-consuming process involving a lot of human effort and is therefore not carried out as often as it should be. Our aim was therefore to explore the potential of machine learning methods to reduce the human workload, and to particularly also gauge the performance of deep learning methods as compared to more established machine learning methods.

**Methods:** We used three available reviews of diagnostic test studies as data basis. In order to identify relevant publications we used typical text pre-processing methods. The reference standard for the evaluation was the human-consensus based binary classification (inclusion, exclusion). For the evaluation of models various scenarios were generated using a grid of combinations of data preprocessing steps. Furthermore, we evaluated each machine learning approach with an approach-specific predefined grid of tuning parameters using the Brier score metric.

**Results:** The best performance was obtained with an ensemble method for two of the reviews, and by a deep learning approach for the other review. Yet, the final performance of approaches is seen to strongly depend on data preparation. Overall, machine learning methods provided reasonable classification.

**Conclusion:** It seems possible to reduce the human workload in updating systematic reviews by using machine learning methods. Yet, as the influence of data preprocessing on the final performance seems to be at least as important as choosing the specific machine learning approach, users should not blindly expect good performance just by using approaches from a popular class, such as deep learning.

## 1 Introduction

In patient-centered medicine, the integration of external evidence is a crucial component in deciding on the use of medical services. Clinicians, researchers and health policy makers have to deal with a multitude of publications within their field of expertise. A systematic review summarizes the empirical evidence according to a priori defined inclusion criteria into question and serves as an external evidence basis for clinical decisions.^1^ An increase in the number of studies, such as seen in medicine in recent years,^2^ also leads to the need for a higher frequency of systematic review updates. The updating of systematic reviews is very resource-intensive, which creates a major barrier for up-to-date evidence syntheses. Intelligent technical support systems, e.g., based on machine learning techniques, are a promising approach for reducing the human effort in this update process. In particular, deep learning techniques, which have recently become quite popular, are expected to considerably advance this area. We specifically consider the setting of systematic reviews of diagnostic test studies and compare the performance of several machine learning techniques, also including deep learning approaches.

In the context of systematic reviews and living reviews, i.e. continuous updates, various computer-assisted approaches were introduced to increase the efficiency^3-5^. Currently, however, living reviews are not widely used^5^, as the human effort for continuous updating within a few months is very substantial. Despite existing efforts to automate sub-processes of systematic/living reviews, these are only rarely applied in research practice. Initially, studies investigated the performance of established machine learning methods, such as support vector machines or random forests. The results obtained with these have already highlighted the potential of such support systems.^3^ Recently, the application of deep learning methods for natural language processing has been growing in addition to conventional machine learning methods.^6^ Machine learning and in particular deep learning have mostly been successful in big data applications. Here we investigate to what extent such techniques can be useful in settings with relatively small data sizes.

According to our knowledge, only one study by Marshall, Noel-Storr, Kuiper, Thomas and Wallace ^7^ compares convolutional neural networks (CNN) with other machine learning methods in the context of systematic reviews. Furthermore, current research has been mainly focused on the classification in the setting of highly specific reporting standards, such as randomized controlled trials (RCT)s.

We specifically evaluate the performance of machine learning methods for identifying relevant publications in the context of updating a systematic review of diagnostic studies, which are characterized by rather low standardization of reporting, making classification more difficult. In addition to comparing different machine learning techniques, we also focus particularly on the effects of data preprocessing and tuning parameter selection on classification performance, as these might have considerably influence.

In the following we describe the three systematic reviews used for evaluation, the approaches for data processing, the training of machine learning techniques, and evaluation criteria, before reporting results, discussing these and giving some concluding remarks.

## 2 Materials and methods

### 2.1 Data sets

Three reviews were used to assess the performance of various machine learning methods in the context of systematic reviews (Table 1). These are data from the screening process of two already published reviews (R-1^8-10^, R-3^10^) of diagnostic studies in the field of orthopaedics and physical therapy. The review R-2^11^ contains data of an unpublished Cochrane review. The size of the review is typical for diagnostic test systematic reviews in the field of medicine. The investigated systematic reviews aimed at evaluating the validity and reliability of a physical examination test for structural and functional testing of the anterior and posterior cruciate ligament and shoulder pathologies. Included were studies which (1) examined the desired physical target structure and (2) the quality of the physical tests. The original queries were carried out in the following databases: MEDLINE via PubMed/Ovid, EMBASE via Ovid, AMED, CINAHL, DARE, MEDION, and ARIF.

**Table 1:**
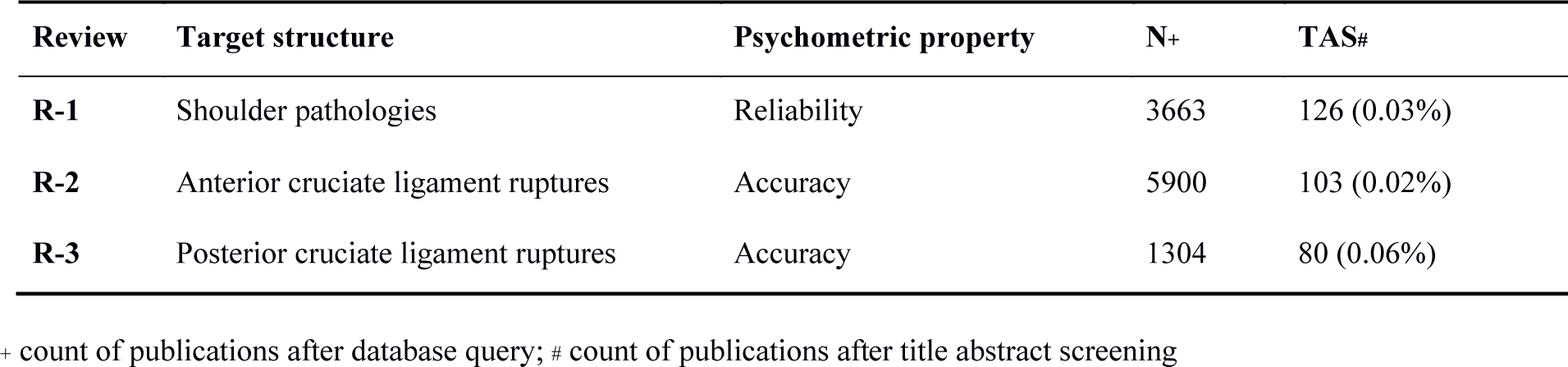
Reviews

| Review | Target structure | Psychometric property | N+ | TAS# |
| --- | --- | --- | --- | --- |
| R-1 | Shoulder pathologies | Reliability | 3663 | 126 (0.03%) |
| R-2 | Anterior cruciate ligament ruptures | Accuracy | 5900 | 103 (0.02%) |
| R-3 | Posterior cruciate ligament ruptures | Accuracy | 1304 | 80 (0.06%) |

+ count of publications after database query; # count of publications after title abstract screening

### 2.2 Study procedure

**Figure 1** illustrates the entire study process, containing text preprocessing, scenario preparation, training of machine learning methods and evaluation. In order to identify relevant publications, the title and abstract of each publication were available as training data. The reference standard for the evaluation of the machine learning methods was the human-consensus based classification (inclusion, exclusion) of the studies based on title and abstract screening. The various phases of the study are described in more detail below.

**Figure 1:**
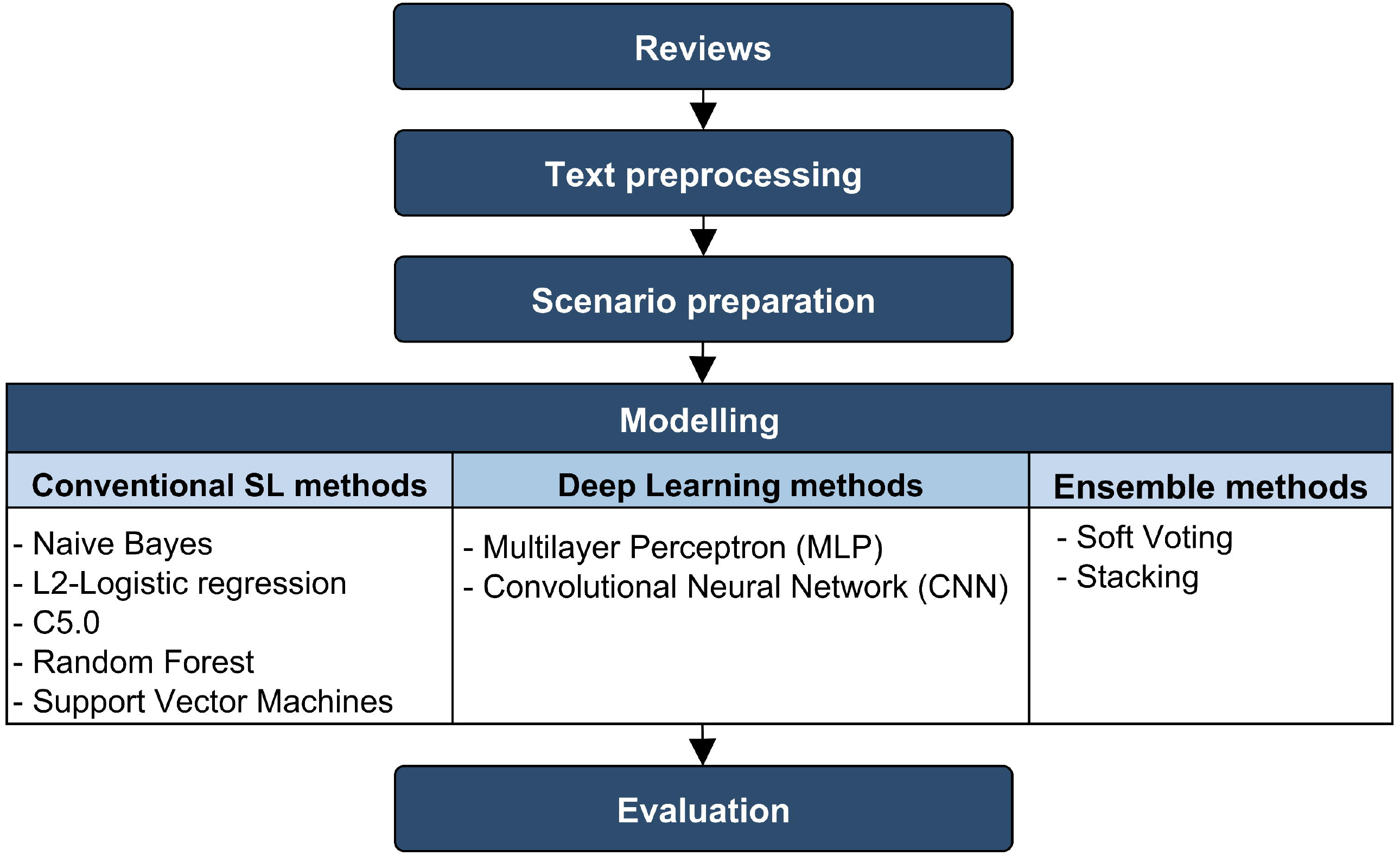
Study procedure

### 2.3 Text preprocessing

After export from the literature databases, the titles and abstracts are available in different quality. Abstracts, for example, varied in structure and sometimes additional unnecessary information were reported and had to be removed. Otherwise, they would have distorted the classification. Therefore, in the first processing step, the texts were preprocessed with the aim of creating a quality-checked data basis and optimizing the information content.

Various text cleaning procedures were used. Additional content was removed using regular expressions in an iterative process (e.g. journal or author information). Furthermore, standard text procedures (punctuation handling and upper/lower case, replacement of numbers and symbols and removal of stopwords) were applied to the reviews. Two different types of text reduction techniques were compared to each other: (1) Stemming - the individual words of the text were reduced to the undeclined root word to identify words with the same content as the same token. (2) Lemmatization - inflected words were grouped together into their original form.

### 2.4 Data scenario preparation

After text preprocessing, the reviews were prepared for model training and various scenarios were generated using a mixture of data preprocessing steps. **Figure 2** outlines this procedure. The aim was to ensure a fixed allocation of publications into training and test data and to provide various data preparation scenarios for model training in a compact way. The sampling ratio of relevant versus irrelevant publications (inclusion/exclusion) was manipulated to investigate a possible effect of the balancing on model training.

**Figure 2:**
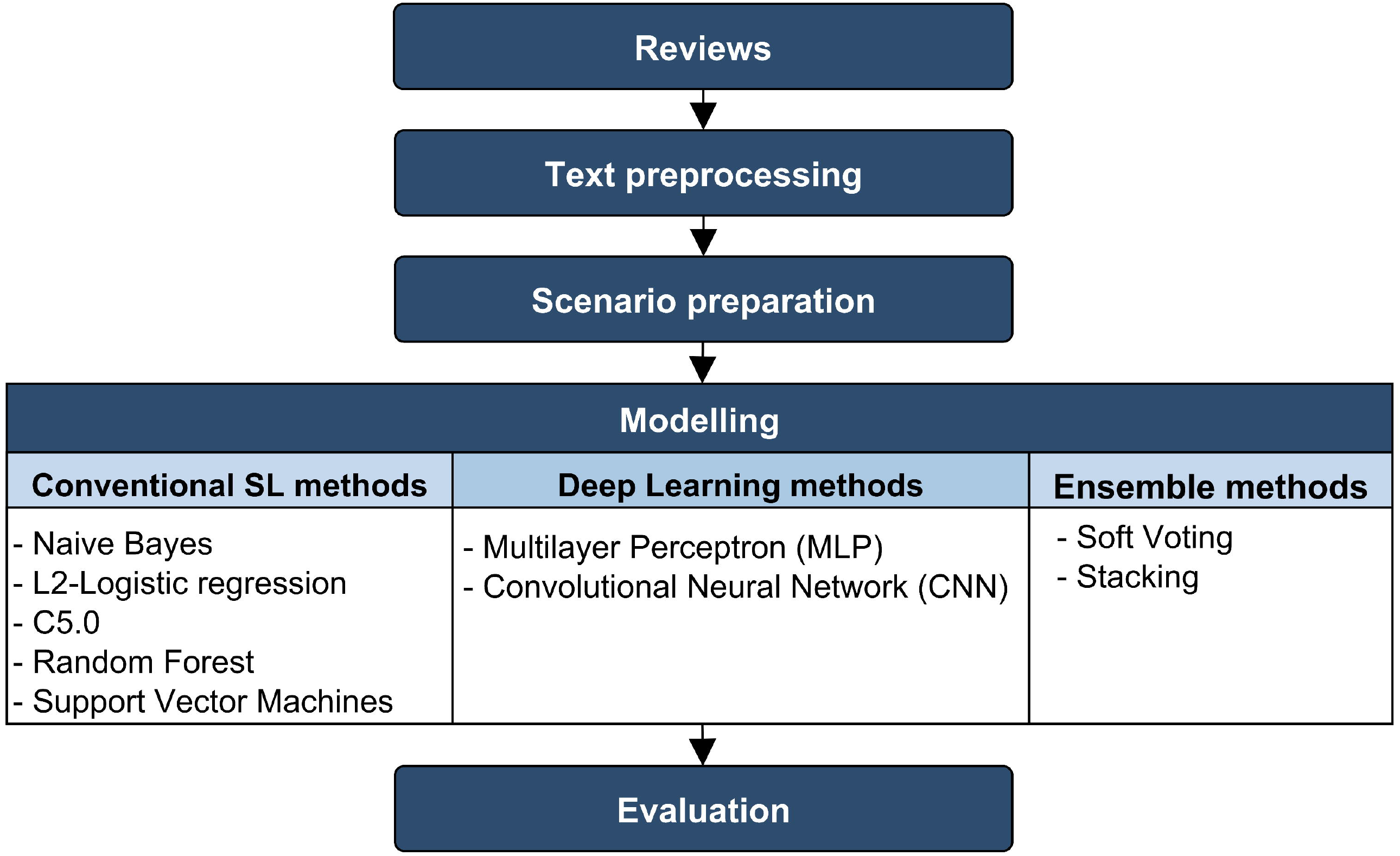
Data preprocessing and scenario preparation

Basically, two different types of data representation were required for the different machine learning methods^12^. On the one hand, (1) the conventional machine learning methods use sparse vectors/document-term-matrices as input data. On the other hand, (2) the DL methods with word embedding require the input data represented as dense vectors. For both types of data representation, we varied (1) the number of n-grams between one and three words/tokens, (2) both types of text reduction techniques and (3) the maximum number of initial tokens (original frequency, fifteen, ten and one thousand and five hundred). Furthermore, for the document-term-matrix, we used the (a) term-frequency (Tf) and (b) the term-frequency – inverse document frequency (Tf-idf) as weighting scheme for the tokens. This results in 150 different data scenarios per review for the conventional machine learning methods and 50 data scenarios for the deep learning approaches.

### 2.5 Word embedding

The word embedding was generated by a skip-gram-model with the preprocessed three reviews as inputs13. In total, two word embeddings were used based on both text reduction techniques (1) stemming and (2) lemmatization. The following parameters were used for the word embeddings: Loss function: Binary crossentropy, Epochs: 10, Batch size 1, Learning rate 0.01, Optimizer: Adam (decacy: 0, *β*_1_: 0.9, *β*_1_: 0.999), Dimensions: 300, Skip window: 5, Negative sample: 1.

### 2.6 Model training

Different models were trained on each specific data scenario (**Figure 2**). Overall, we investigated six conventional machine learning methods, two deep learning methods, and two ensemble methods.

For **conventional machine learning methods:** (SL-1) logistic regression with elastic net regularization, (SL-2) Naïve Bayes, (SL-3) Support Vector Machines, (SL-4) K-Nearest Neighbors, (SL-5) C5.0 trees, and the (SL-6) Random Forest algorithm were used (**Supplement Table 1**).

Further, we investigated the performance of different **deep learning methods:** (DL-1) Multilayer Perceptron (MLP) – Bag of Words approach, (DL-2) MLP – with word embedding, (DL-3) Convolutional Neural Networks (CNN) –with word embedding, (DL-4) CNN-MLP – with word embedding (**Supplement Table 2**).

Two different **ensemble methods**, which comprise conventional machine methods, were evaluated. (E- 1) Ensemble stack with a Random Forest as meta learner and the (E-2) ensemble soft voting, averaging the prediction probabilities of the base learners. Both Ensembles (E-1, E-2) used the scenario specific best trained base learner (B-1) logistic regression with elastic net regularization, or (B-2) Support Vector Machines, as well as (B-3) a Random Forest algorithm. In addition, a soft voting ensemble was used to combine (E-3) deep learning methods.

The pseudo code in **Figure 3** illustrates the entire model training process. For each model, a 10-fold cross-validation with 3 repetitions was used to identify the optimal tuning parameter set for each data scenario. To enable a fair comparison, it was ensured that resampling was identical across all models during cross-validation. The optimal data scenario for each algorithm with its specific optimal tuning parameters was determined by averaging across hold-out predictions while training and choosing the best performing scenario.

**Figure 3:**
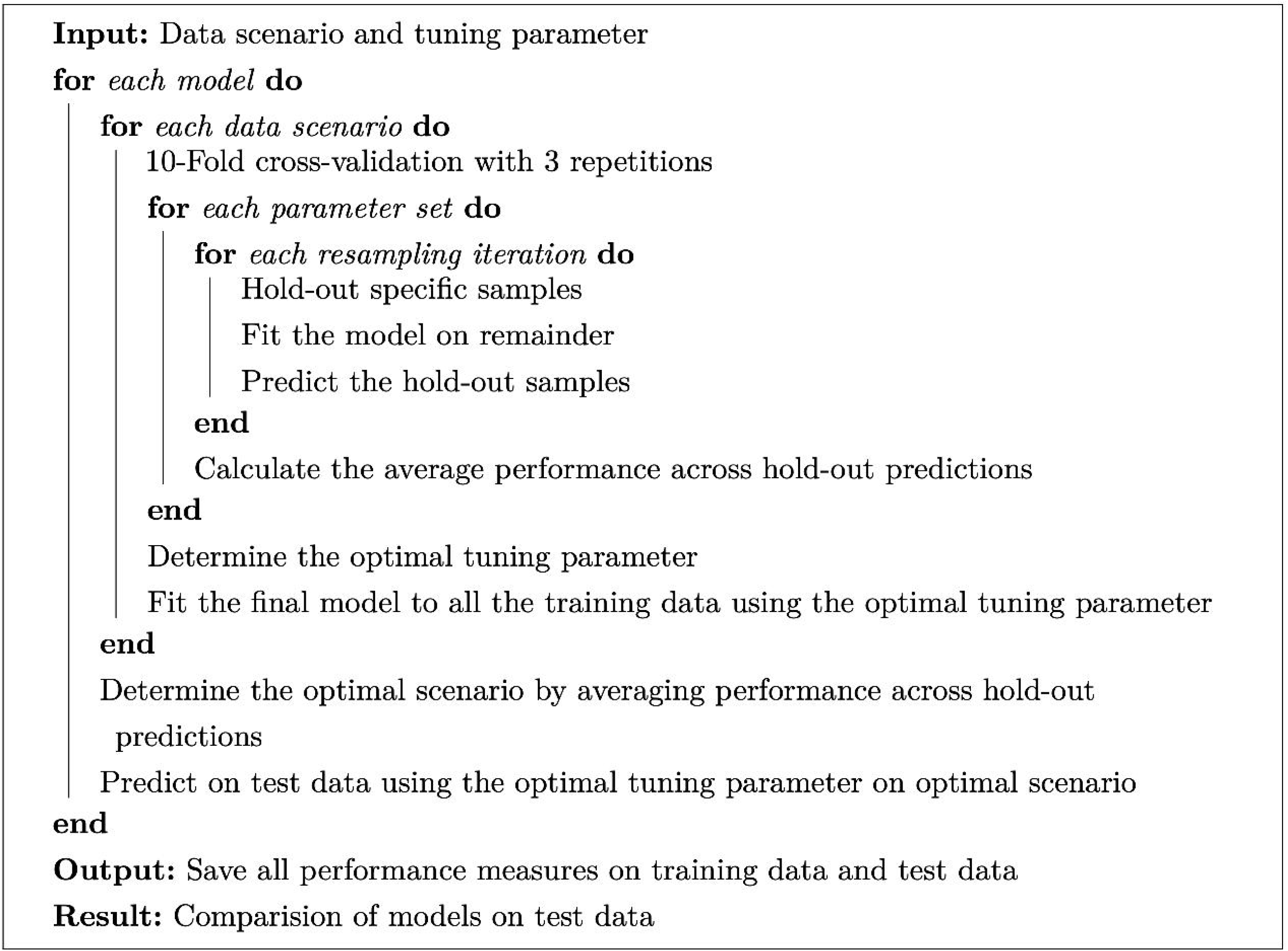
Algorithm: model training and practical implementation of model training; adapted from Kuhn ^14^

### 2.7 Model evaluation

The evaluation of the methods was carried out step by step. As outlined in **Figure 3**, (1) the optimal tuning parameter constellation was selected per scenario, (2) the optimal scenario per method was identified, and then (3) the optimal tuning parameters per method and scenario were used to determine the predictive value of the respective method on the independent test data.

1. The Brier score (Formula 1) was primarily used as the selection criterion for the optimal tuning parameters. This score describes the precision of the predictions by determining the distance of the predicted probabilities from the reference standard^15^.
2. To enable a fair comparison between the scenarios with differently balanced proportions, the Brier score for each scenario was compared to the scenario-specific null model. We call this “Brier comparison score” and used this score to determine the optimal scenario (Formula 2). As null model we trained an intercept only model.
3. After the models were trained with the optimal tuning parameter set on the optimal scenario, the final prediction probabilities were determined on a corresponding independent test data set. The Brier comparison score was primarily used as evaluation metric. Other metrics, such as sensitivity, specificity, as well as positive and negative predictive values, were also calculated.

Formula 1: Brier score

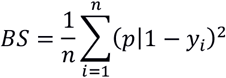

Formula 2: Brier comparison score

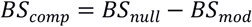

### 2.8 Sensitivity analysis

To compensate for the high class imbalance (few inclusions/many exclusions) that is inherent in all title and abstract screenings, we lowered the default threshold value (50:50) for binary classification to evaluate its influence on the prediction success. The threshold was reduced from 0.5 to 0.2 to improve the classification sensitivity of the models (i.e. lower the false negative rate).

All statistical analyses were performed using the statistical software R^16^. For training and systematic evaluation the Caret package^14^ was used which allows to streamline for creating predictive models including the implementation of self-developed algorithms, loss functions, and evaluation metrics. This package also enables the evaluation of the models in direct comparison in a sufficient manner.

## 3 Results

Overall, three different but thematically similar systematic medical reviews were analyzed. Specifically, only titles and abstracts were used and no full texts. An abstract contains on average 225.2 ± 117.4 words (median= 226; min/max= 2/708). After data preprocessing (stemming and lemmatization) the maximum overall number of tokens (words) per review varied within a range of: R-1: 16.807/19.162; R-2: 12.722/14.684; R-3: 5.924/6.905.

The data preprocessing grid (shown in **Figure 2**) resulted in 1038 different data scenarios in total (Document Term Matrix – R-1: 396; R-2: 342; R-3: 300; Dense-Vector representation – R-1: 66; R-2: 57; R-3: 50).

### 3.1 Evaluation of data scenarios

**Figure 4** gives an overview of model comparison during the training step (cross validation) for one review (R-1) as example. Each data point represents a prediction for a specific data preparation scenario as measured by the Brier score. On the y-axis the Brier comparison score is shown. Positive values indicate a better performance of the model than the scenario specific null model, which is based on an intercept only model. Note that some methods tend to result in rather poor predictions, with negative Brier comparison scores (e.g. K-NN, Naïve Bayes). Furthermore, variation between models within one algorithm is higher than the variation between different algorithms. Depending on the specific data preparation procedures, a specific method might perform well or rather poor. Yet, there is no clear relationship between the specific data preparation steps and the performance of the methods. The algorithms perform differently on the same data preparation scenarios, but with no distinguishable patterns. It seems that each algorithm needs its own optimal preprocessing steps, also strongly depending on the input data. Two different data preprocessing methods that were designed to reduce the dimensionality of the feature space show impact on model performance in many cases. One is token reduction, as a method to increase the signal to noise ratio, another are *word embeddings*, which are specific to deep neural networks. *Word embeddings* take into account the proximity of words and generate a lower dimensional vector out of a high dimensional matrix. It can then be used to pre-weight the input data of a deep neural network. The above findings remain consistent across all reviews examined.

**Figure 4:**
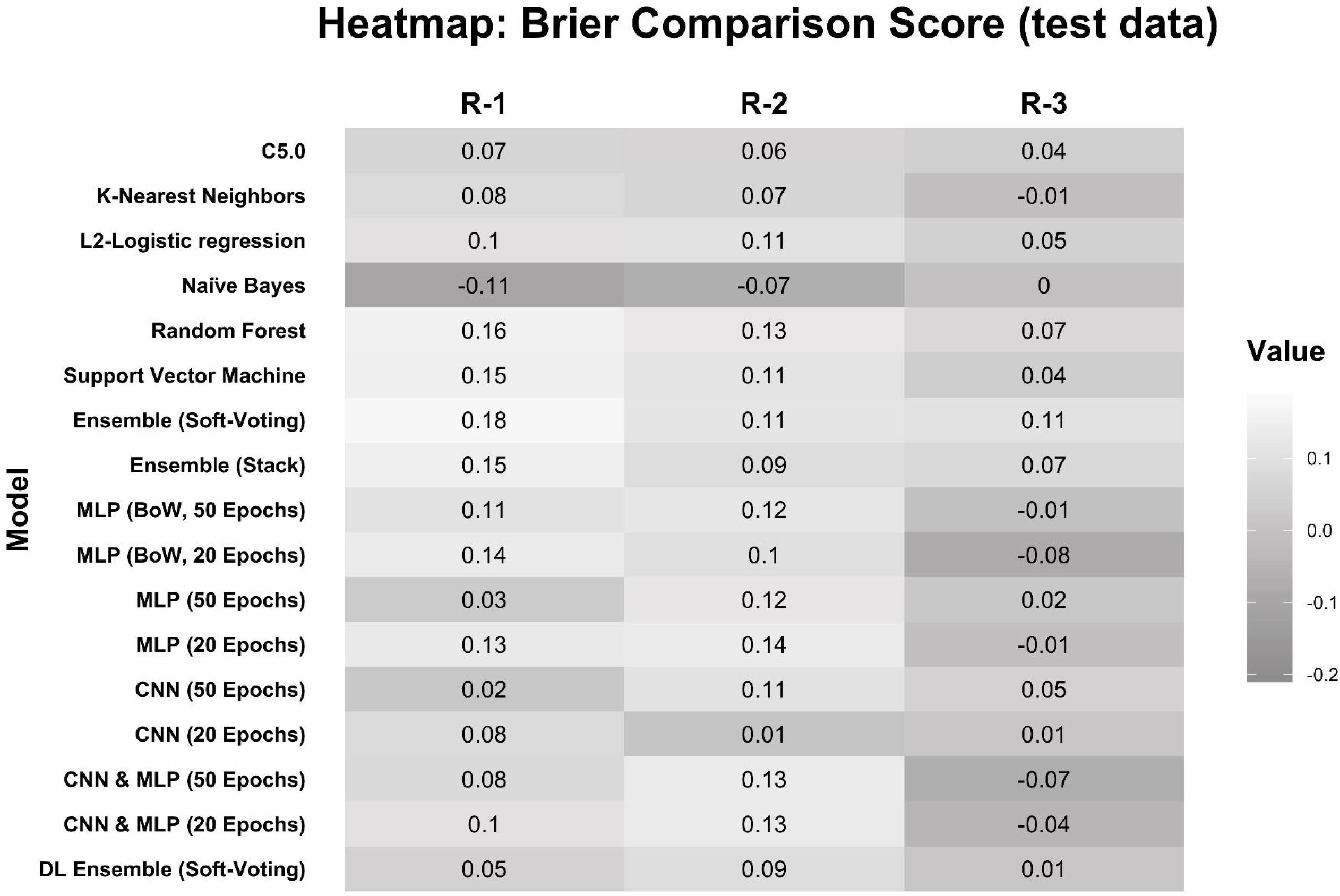
Comparison of data scenarios

### 3.2 Evaluation of test data performance

The final evaluation of model performance (i.e. generalizability) is based on independent test data. **Figure 5** shows a heatmap with Brier comparison scores for all investigated reviews (R1-3). Cells with light gray background (values above zero) point to a better performance of the corresponding model compared to the scenario specific null model, while dark cells (values below zero) indicate worse performance. The best performing algorithm varies between the reviews examined (R1: Ensemble (Soft-Voting), R2: MLP (20 epochs), R3: Ensemble (Soft-Voting)). Across all reviews, there is no clear winner and the performance varies between the reviews. Review R3 is most challenging during test data prediction. In review R2, deep learning outperforms most conventional machine learning methods, while in review R1 and R3 ensembles of conventional machine learning methods are superior.

**Figure 5:**
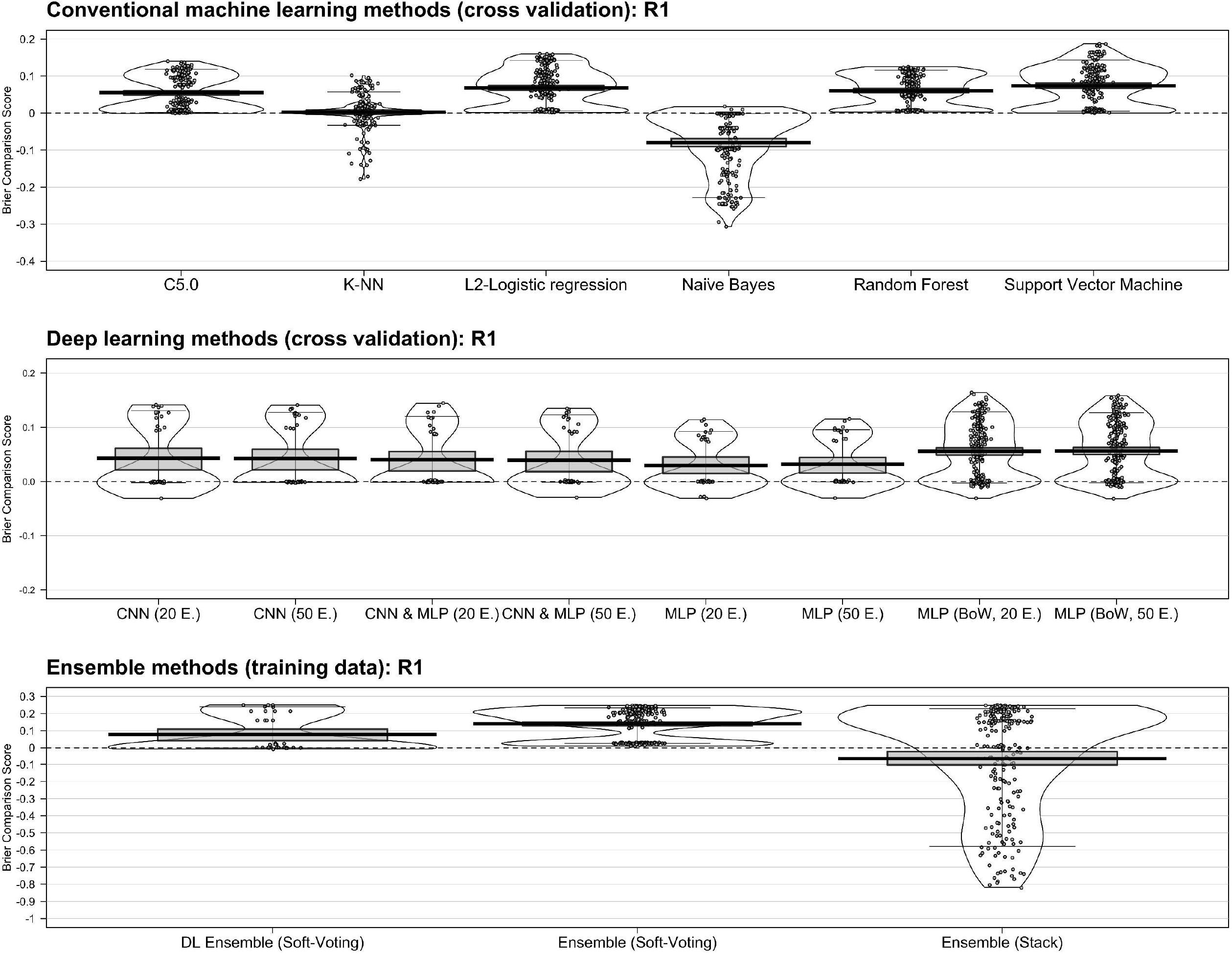
Heatmap – performance on unseen test data based on the Brier comparison score

Note that not all possibilities of model tuning, especially *thresholding* or generalized *word embeddings*, were fully systematically investigated for deep learning methods. Depending on the evaluation metric, the best performing algorithm changes within a review. In the present case of updates of systematic reviews, there is always a strong class imbalance, with the minority class being of particular interest. Under this scenario a stable evaluation metric is needed incorporating this high class imbalance. This would certainly improve the quality of the results. Ensemble methods, incorporating more than one algorithm, may lead to better results here compared to single algorithms since multi-model ensembles increase the sensitivity of the analysis purely by incorporating more than one algorithm, especially by aggregating with plurality or soft voting.

### 3.3 Sensitivity analysis

For sensitivity analysis, we lowered the classification threshold from 0.5 (equal probabilities on both sides) to 0.2 inducing a more sensitive classification. This clearly leads to better results since a more sensitive classification is superior in this task. Not missing a potentially relevant manuscript and therefore increasing the inclusion rate (lower the false negative rate) is of advantage here (**Supplement Table 3, Supplement Table 4, Supplement Table 5**).

## 4 Discussion

We investigated the performance of machine learning methods for systematic reviews in the challenging setting of diagnostic test studies. Besides the general level of performance, we were interested whether deep learning approaches would have a considerable advantage of conventional machine learning methods. We found that deep neural network approaches were among the top performers but did not considerably outperform conventional machine learning methods such as random forest or ensembles. This might be due to the rather low number of publications and the rather small amount of text available from titles and abstracts.

Yet, searching for the best algorithm for the task is only part of the picture. Instead our results suggest that data preprocessing steps had much more influence on the final performance. With a bad choice of preprocessing steps any approach could drop to or even below the performance level of the scenario specific null model, which only uses an intercept as predictor variable and no words. On the other hand, most algorithms could perform rather well on a specific combination of data preprocessing steps. Unfortunately, it seems difficult to predict in advance which preprocessing steps are best suited for which algorithm and review. Therefore, we strongly encourage to consider data preprocessing besides other predictors as a tuning parameter for training of machine learning approaches. Yet, one consistent pattern is that technics that reducing the dimensional load of the models are preferable. For example, token reduction, which increases the signal to noise ratio, almost always is beneficial. The same trend is observed for embeddings which reduce dimensions in deep neural network models.

In the process of updating systematic reviews it is of utmost importance not to miss a relevant publication, thus high sensitivity is needed. In fact, in our medical context we want to achieve a sensitivity of one. Accordingly, we required that a usable algorithm must have a sensitivity of one and a specificity that allows to reduce the number of texts to be examined manually at least by a factor of two. By lowering the binary classification threshold from equal probabilities to 0.2 we were able to achieve such results with a sensitivity of one. At the same time, our goal of automatically sorting out at least 50 percent of all texts was achieved in all three reviews. However, we did not use a systematic search for the optimal threshold for each review, instead freely chose a value of 0.2 for all sensitivity analyses. A separate test data set would in principle allow cross-validation to find the optimal threshold for the specific problem.

### 4.1 Methodological considerations

A stable comparison metric in the setting of high class imbalance is desirable to clarify there the question which algorithm works best. In the presence of a high class imbalance, all used evaluation metrics, including the Brier comparison score, the null models tend to be extremely good performing. In fact, the higher the imbalance the better the null model. Another potential measure is the Net Benefit^17^, which considers the class imbalance by weighting the difference in true and false positive rate. However, the advantage of the Brier score is the incorporation of the distance of the predicted probabilities. In contrast, the Net Benefit uses only the binary classification. Thus, an inclusion of a prevalence adjusting weighting factor for the Brier score could improve the performance in class imbalance problems. Maybe, a combination of various metadata could increase the classification performance. In addition to title and abstract, further information such as full text, MeSH term indexing or hierarchical inclusion of keywords amongst others could be used. Thus, separate classifiers trained on the different input information could be combined for improved predictions e.g. with a majority-vote ensemble.

### 4.2 Strengths and weaknesses

One of the main strengths of this work is the realization and implementation of all working processes in a unified development environment, which allow us to systematically generate and evaluate different models on different data sets with different data pre-processing steps. According to the review by O’Mara-Eves, Thomas, McNaught, Miwa and Ananiadou ^3^ the Brier Score, was not included in any recent scientific research of machine learning methods as evaluation metric in the context of updating systematic reviews. In favor of a balanced prevalence in training and test data sets, a putative interesting time effect was not considered (e.g. changes in quantity and structure in reporting over time). The tested machine learning and deep learning methods represent only a selection of the methods potentially available for this classification task.

## 5 Conclusions

Overall, the presented work shows that it is possible to reduce the human work load in updating scientific systematic reviews by machine learning methods, even for studies with a low standardized reporting quality. There is no big performance difference between deep learning and other machine learning approaches. Instead, the influence of data preprocessing on the final model performance is rather strong. Optimizing the threshold assessment and evaluating additional, desirable stable, metrics in high class imbalance settings seems more urgent and should be the topic of future research.

## 7 Declarations

### 7.1 Ethics approval and consent to participate

Not applicable

### 7.2 Consent for publication

Not applicable

### 7.3 Availability of data and material

The data sets used and/or analyzed during the current study are available from the corresponding author on reasonable request.

### 7.6 Authors’ contributions

Author Contribution: TL, HB, GS, TD jointly conceived the study. TL performed the analysis. All authors drafted the paper and have critically revised it and approved the final version.

## 7.7 Acknowledgements

We would like to thank Christian Kopkow and Alice Freiberg for providing their screening data for model training.

## 8 Supplement

**Supplement Table 1:** Tuning parameter of conventional machine learning methods

| Model | R package | Parameter | Grid |
| --- | --- | --- | --- |
| **Naïve Bayes** | naivebayes | usekernel | [TRUE, FALSE] |
|  |  | laplace | [0, 1] |
|  |  | adjust | [0, 1, 2] |
| **Logistic regression** | glmnet | alpha | [seq(from = 0, to = 1, length.out = 10)] |
|  |  | lambda | [10^seq(from = 3, to = -3, length.out = 100)] |
| **C5.0** | C50 | trials | [1:9, 10:100] |
|  |  | model | [tree, rules] |
|  |  | winnow | [TRUE, FALSE] |
| **Random Forest** | ranger | mtry | [sqrt(ncol(data) - 1)] |
|  |  | Splitrule | [gini, extratrees] |
| **Support Vector Machine** | e1071 | cost | [0.2, 0.5, 1, 5, 10, 30, 60] |
|  |  | weight | [seq(0.5, 1.5, 0.1)] |
| **K-Nearest Neighbors** | kkn | k | [3, 6, 9, 12, 15, 18] |
| **Ensemble (Stack)** | caretEnsemble | rf | - |
| **Ensemble (Soft-Voting)** | - | - | - |

**Supplement Table 2:** Deep learning methods - Network architecture

| Global Parameter | MLP (Bag of Words) | MLP (Word embedding) | CNN (1) | CNN (2) |
| --- | --- | --- | --- | --- |
| **Loss-Function** | Binary cross entropy | Binary cross entropy | Binary cross entropy | Binary cross entropy |
| **Epochs** | [20, 50] | [20, 50] | [20, 50] | [20, 50] |
| **Batch size:** | 20 | 20 | 20 | 20 |
| **Learning rate** | 0.01 | 0.01 | 0.01 | 0.01 |
| **Optimizer** | Adam | Adam | Adam | Adam |
| **Architecture** | Fully-connected-Layer 1 (units: 512, activation: ReLU, dropout: 0.2)<br>Fully-connected-Layer 2 (units: 256, activation: ReLU, dropout: 0.2)<br>Fully-connected-Layer 3 (units: 64, activation: ReLU)<br>Fully-connected-Layer 4 (units: 2, activation: softmax) | Word embedding-Layer<br>Global-Max-Pooling-Layer<br>Fully-connected-Layer 1 (units: 300, activation: ReLU)<br>Fully-connected-Layer 2 (units: 128, activation: ReLU)<br>Fully-connected-Layer 3 (units: 32, activation: ReLU)<br>Fully-connected-Layer 4 (units: 2, activation: softmax) | Word embedding-Layer<br>Convolutional-Layer (filter: 150, kernel size: 3, activation: ReLU)<br>Global-Max-Pooling-Layer<br>Fully-connected-Layer 2 (units: 2, activation: softmax) | Word embedding-Layer<br>Convolutional-Layer (filter: 150, kernel size: 3, activation: ReLU)<br>Global-Max-Pooling-Layer<br>Fully-connected-Layer 1 (units: 32, activation: ReLU)<br>Fully-connected-Layer 2 (units: 2, activation: softmax) |

\* All DL-methods used callbacks (1) early stopping after 10 epochs with no further increasing of accuracy; (2) reduce learning rate on plateau with patience 5 effectively reduced the learning rate by 10%.

**Supplement Table 3:**
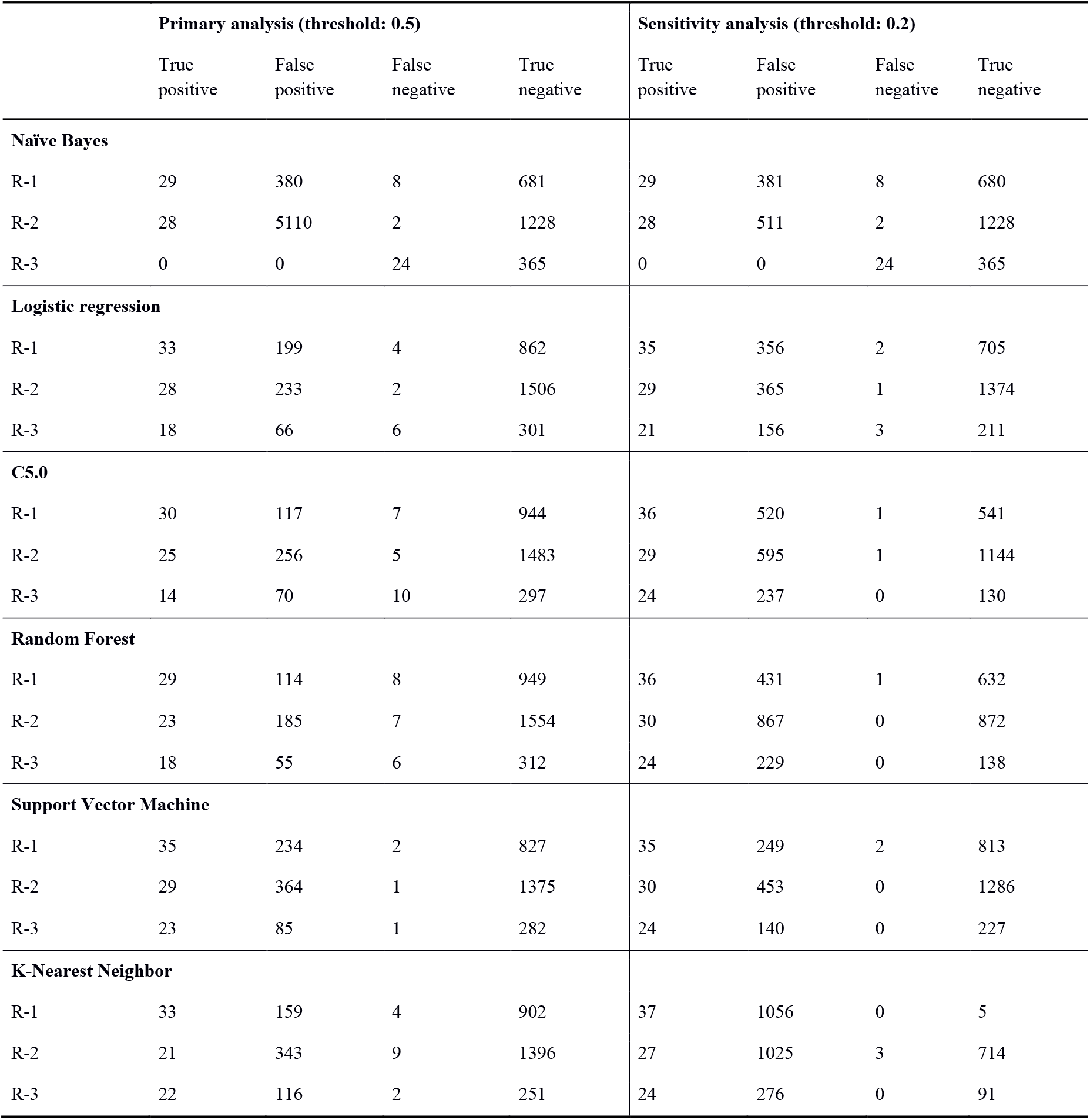
2×2 table – conventional machine learning methods

**Supplement Table 4:** 2×2 table – Deep learning methods

|  | Primary analysis (threshold: 0.5) |  |  |  | Sensitivity analysis (threshold: 0.2) |  |  |  |
| --- | --- | --- | --- | --- | --- | --- | --- | --- |
|  | True positive | False positive | False negative | True negative | True positive | False positive | False negative | True negative |
| **MLP (BoW, 50 Epochs)** |  |  |  |  |  |  |  |  |
| R-1 | 33 | 156 | 4 | 905 | 33 | 164 | 4 | 897 |
| R-2 | 27 | 184 | 3 | 1555 | 27 | 191 | 3 | 1548 |
| R-3 | 20 | 79 | 4 | 288 | 20 | 84 | 4 | 283 |
| **MLP (BoW, 20 Epochs)** |  |  |  |  |  |  |  |  |
| R-1 | 32 | 121 | 5 | 940 | 33 | 183 | 4 | 878 |
| R-2 | 27 | 223 | 3 | 1516 | 24 | 202 | 6 | 1537 |
| R-3 | 21 | 110 | 3 | 257 | 11 | 17 | 13 | 350 |
| **MLP (50 Epochs)** |  |  |  |  |  |  |  |  |
| R-1 | 33 | 161 | 4 | 900 | 31 | 171 | 6 | 890 |
| R-2 | 24 | 184 | 6 | 1555 | 22 | 201 | 8 | 1538 |
| R-3 | 11 | 13 | 13 | 354 | 11 | 17 | 13 | 350 |
| **MLP (20 Epochs)** |  |  |  |  |  |  |  |  |
| R-1 | 27 | 56 | 10 | 1005 | 34 | 354 | 3 | 707 |
| R-2 | 25 | 161 | 5 | 1578 | 27 | 304 | 3 | 1435 |
| R-3 | 14 | 30 | 10 | 337 | 21 | 96 | 3 | 271 |
| **CNN (50 Epochs)** |  |  |  |  |  |  |  |  |
| R-1 | 33 | 285 | 4 | 776 | 33 | 164 | 4 | 897 |
| R-2 | 26 | 238 | 4 | 1501 | 27 | 210 | 3 | 1529 |
| R-3 | 20 | 61 | 4 | 306 | 19 | 58 | 5 | 309 |
| **CNN (20 Epochs)** |  |  |  |  |  |  |  |  |
| R-1 | 32 | 215 | 5 | 847 | 33 | 269 | 4 | 792 |
| R-2 | 28 | 432 | 2 | 1307 | 19 | 122 | 11 | 1617 |
| R-3 | 21 | 80 | 3 | 287 | 11 | 17 | 13 | 350 |
| **CNN & MLP (50 Epochs)** |  |  |  |  |  |  |  |  |
| R-1 | 32 | 214 | 5 | 847 | 32 | 208 | 5 | 853 |
| R-2 | 28 | 432 | 2 | 1307 | 22 | 201 | 8 | 1538 |
| R-3 | 21 | 80 | 3 | 287 | 22 | 135 | 2 | 231 |
| **CNN & MLP (20 Epochs)** |  |  |  |  |  |  |  |  |
| R-1 | 30 | 77 | 7 | 984 | 34 | 354 | 3 | 707 |
| R-2 | 23 | 173 | 7 | 1566 | 29 | 351 | 1 | 1388 |
| R-3 | 20 | 106 | 4 | 261 | 21 | 96 | 3 | 271 |

**Supplement Table 5:** 2×2 table – Ensemble methods

|  | Primary analysis (threshold: 0.5) |  |  |  | Sensitivity analysis (threshold: 0.2) |  |  |  |
| --- | --- | --- | --- | --- | --- | --- | --- | --- |
|  | True positive | False positive | False negative | True negative | True positive | False positive | False negative | True negative |
| **Ensemble (Stack)** |  |  |  |  |  |  |  |  |
| R-1 | 32 | 152 | 5 | 911 | 34 | 222 | 3 | 841 |
| R-2 | 24 | 171 | 6 | 1567 | 26 | 305 | 4 | 1433 |
| R-3 | 17 | 62 | 7 | 305 | 24 | 126 | 0 | 241 |
| **Ensemble (Soft-Voting)** |  |  |  |  |  |  |  |  |
| R-1 | 29 | 81 | 8 | 982 | 34 | 275 | 3 | 788 |
| R-2 | 27 | 242 | 3 | 1497 | 30 | 587 | 0 | 1152 |
| R-3 | 17 | 45 | 7 | 322 | 24 | 142 | 0 | 225 |
| **DL Ensemble (Soft-Voting)** |  |  |  |  |  |  |  |  |
| R-1 | 32 | 273 | 5 | 788 | 35 | 390 | 2 | 671 |
| R-2 | 29 | 266 | 1 | 1473 | 29 | 438 | 1 | 1301 |
| R-3 | 19 | 89 | 5 | 278 | 22 | 132 | 2 | 235 |

